# 40HZ photoacoustic interventions combined with blue light improve brain function and sleep quality in a healthy population

**DOI:** 10.1101/2023.05.22.23290180

**Authors:** Xiaojun Xu, Zhuping Gong, Kaixiu Jin, Peiyu Huang, Cheng Zhou, Qingze Zeng, Junhu Teng, Ziyu Liu, Huajiang Zhang, Qian Xu, Yanling Chen, Cong Fang, Minming Zhang

## Abstract

Recent studies suggest that 40Hz photoacoustic stimulation is beneficial for improving cognition, and is safe and effective in patients with Alzheimer’s disease (AD) and mild cognitive impairment (MCI). It has been reported that the healthy population responds more strongly to the 40 Hz intervention compared to MCI and AD patients. Therefore, the aim of this study was to explore the effectiveness of 40 Hz photoacoustics in preventing cognitive impairment and improving sleep quality in the healthy population.

Four weeks of 40 Hz intervention significantly improved sleep quality (PSQI) (p=0.028) and enhanced functional connectivity in the hippocampus and default mode network (DMN) (in the cognitively impaired population, functional connectivity showed a decreasing trend). These results were consistent with findings from the FDA clinical trial on MCI/AD patients that showed enhanced connectivity of the core regions PCC and mPFC within the DMN network. No serious adverse effects were reported throughout the intervention.

This study is the first to validate the efficacy and safety of a 40 Hz photoacoustic intervention in a healthy population, suggesting its potential in preventing cognitive decline in a healthy population.

## Introduction

Aging mainly causes brain atrophy and decline in brain function. Increase in the aging population is a global challenge that has led to the significant increase in the burden of neurological disorders (Wang Y, 2020). According to the Global Burden of Disease Study, neurological disorders are the leading cause of disability-adjusted life years (276 million) globally, and the second leading cause of death in 2016 accounting for 9 million deaths (GBD 2019). It is estimated that by 2030, more than 82 million people worldwide will suffer from cognitive impairment, with health care costs reaching $2.54 trillion (Jia J, 2018).

The health condition of the brain in the healthy population is poor even before the onset of clinical symptoms. Unhealthy lifestyles such as smoking and drinking, lack of physical activity, irregular diet, late nights and stress (Xu W, 2015) and increase in incidences of chronic diseases such as diabetes and hypertension directly or indirectly contribute to decreasing brain function at younger ages and leads to increased brain damage (Pan Y, 2020). Decline in quality of sleep is one of the important behavioural factors affecting brain health (Girardeau G, 2021). A study involving 10 countries worldwide (N=35,327) found that 24% of subjects reported that they did not sleep well. According to the AIS self-assessment, 31.6% of subjects had ‘insomnia’, while 17.5% were considered to have ‘subthreshold insomnia’. ESS scores indicated that 11.6% of subjects were ‘very sleepy’ or ‘dangerously sleepy’ during the day, while in China, up to 45.4% of respondents had experienced varying degrees of insomnia in the preceding month (Soldatos CR. 2005).

Currently, there are no safe and effective strategies of preventing and delaying dementia and other causes of cognitive decline in the brain, including Alzheimer’s disease, Parkinson’s, stroke, and encephalitis (Tisher A, 2019). Non-pharmacological therapies such as photoacoustic intervention (He Q, 2021), electro/magnetic therapy (Chang CH, 2018), cognitive games (Sabbagh M, 2020), and aerobic exercise (Gaitán JM, 2021) have been widely explored to slow the progression of brain cognition-related disorders and improve brain cognition. In 2016, an MIT team found that non-invasive 40Hz light intervention reduced A β1-40 and A β1-42 levels in the visual cortex of AD model mice and reduced plaque load in mice (Laccarino HF, 2016). In 2019, the same team demonstrated that 40Hz light intervention significantly improved neuronal degeneration, improved synaptic function, enhanced neuroprotective factors and reduced DNA damage in neurons of different models of mice, while also reducing the inflammatory response of microglia (Adaikkan C, 2019). More surprisingly, data from human studies conducted in 2021 further validated the effect of this regiment on sleep and cognitive function in AD and MCI patients (He Q, 2021. Cimenser A, 2021).

So far, studies on 40Hz photoacoustic interventions have focused on subjects with cognitive decline related to AD, and little has been reported on the use of this technology in healthy people with poor sleep quality or memory loss. Previous studies of 40Hz interventions in people with AD or MCI reported that the intervention needs to be carried out for 1 hour per day for 3-6 months (Cimenser A, 2021) for improvements in cognitive function and sleep to be observed. Studies have also shown that the same intensity of 40Hz photoacoustic intervention has a higher EEG response in healthy people than in sick people (Chan D, 2022). The aim of this study was to explore the effectiveness and safety of combining 40Hz acoustic intervention with blue light for a 1-month period in improving brain function and sleep quality in a healthy population. We also assessed if the multi-light intervention can shorten the effective intervention time.

## Methods and Materials

### Study design

We first used EEG to evaluate the effect of 5 stimulation modalities of 40Hz photoacoustic stimulation on the response rate of the whole brain 40Hz Gamma oscillations in a healthy population. The five stimulation modalities employed in this study were light-only stimulation (visual stimulation by looking straight at the device and visual stimulation by looking at the front horizontally), audio-only stimulation, and combined stimulation modalities (combination of the two visual stimulation methods with audio stimulation). The aim of this experiment was to determine the optimal stimulation modality for the use of 40 Hz photoacoustic stimulation in the healthy population. The optimal stimulation modality should strike a balance between high response rate of Gamma oscillation generated in the subjects and high tolerance that helps the subjects sustain the daily use of the stimulation device.

Based on the optimal stimulation modality identified from the above experiment, potential subjects were further assessed for sleep quality using the PSQI scale. The effect of 4-week 40Hz photoacoustic intervention combined with blue light on brain function and sleep quality improvement was explored in healthy subjects in need of sleep improvement. All subjects received similar intervention at the same time, from 10:00am-12:00pm each day Monday-Friday, consisting of 30 minutes of blue light stimulation followed by 2 rounds of 30 minutes of 40Hz photoacoustic stimulation, with a 5 minute break after every 30 minutes.

Subjects underwent EEG assessment and fMRI assessment at baseline. MRI scans were performed using Shanghai United Imaging uMR790-3.0T exploratory MRI with a package of T1 structural images, diffusion tensor imaging, resting state functional MRI and arterial spin labeling imaging with EEG use. The main indicators of the study were the improvement in PSQI scores and changes in resting-state functional connectivity (rs-FC), and the safety and compliance of device use. Safety was assessed using an adverse event questionnaire completed during on-site surveys with subjects once a week and an evaluation of abnormal episodes of stimulus response. Adherence was assessed by recording the time each subject used the device at the intervention site.

All participants gave written consent and the study was approved by the Ethics Committee of the Second Affiliated Hospital of Zhejiang University.

### Audiovisual device

The device used for 40Hz photoacoustic plus blue light stimulation in this study was produced by Hangzhou Shudan Medical Technology Co. The blue light of the device consisted of visible light with an average light intensity of 300lux at a wavelength of 464-470nm. The auditory stimulus at 40Hz was a 60dB sound with a 1ms rectangular pulse at every 25ms interval (4% duty cycle, 24ms isi). The white light stimulus at 40Hz consisted of an average light intensity of 450lux at a wavelength of 460-630nm. The visible light consisted of light that can cover the upper side of the eye (see patent specification, Patent No. ZL202221822557.X and Patent No. 202210071845.4, China).

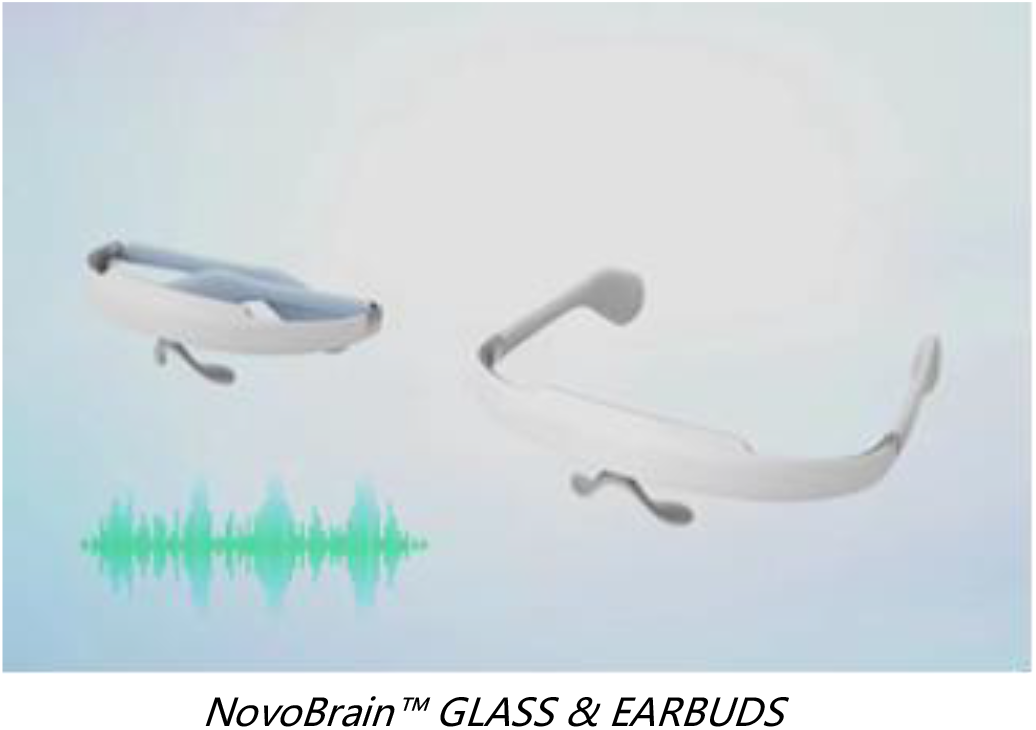

### Participants

Eighteen potential subjects were screened for eligibility in the study. EEG was used to assess the response of this group to 40Hz photoacoustic stimulation under different stimulation modalities. This assessment was used to exclude individuals who were intolerant or did not respond to the stimulation, and then to conduct PSQI evaluation. A total of 6 healthy subjects with need for sleep intervention, were included in this study. See Appendix S1 table for complete inclusion and exclusion criteria. The final number of subjects included in the analysis was 4 since 1 subject was excluded for taking medication to improve insomnia and another subject had severe symptoms of the COVID-19 and therefore did not participate in the subsequent intervention (**Figure 1**).

**Figure 1.**
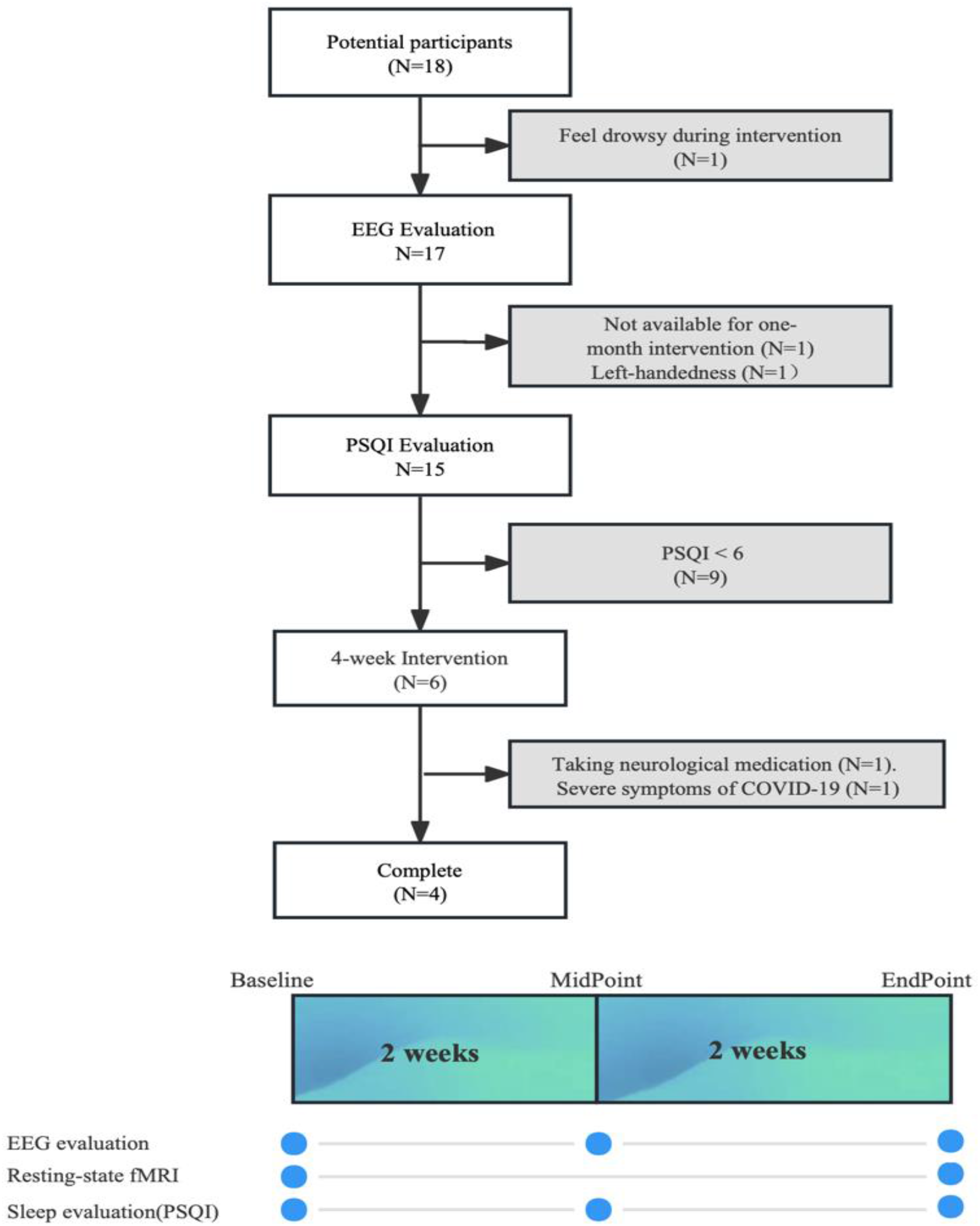
Flow diagram and study design. EEG evaluation was carried out in 17 healthy participants, while only 4 healthy participants received the 4-week intervention. EEG evaluation and PSQI scale were conducted at baseline, the end of 2nd week and the end of 4th week. fMRI were taken at baseline and the end of the 4th week.

### Outcome

The main objective of the study was to explore the safety, stability and effectiveness of a long-acting 40Hz photoacoustic stimulation combined with blue light in improving sleep quality and brain function in healthy subjects.

Safety was assessed using an adverse event questionnaire completed during a weekly on-site survey with participants. Adverse events were tracked during the primary study (4 weeks of 40Hz photoacoustic stimulation combined with blue light), and during follow-up (12 weeks), with an open-label extension underway at the time of publication of this report. Stability of the intervention was assessed by measuring changes in EEG signals from whole brain 40Hz Gamma oscillation stimulated in 4 healthy subjects at 0 week, 2nd week, and 4th week of the optimal stimulation intervention (visual stimulation by looking at the front horizontally+audio stimulation). Sleep quality was also assessed at 0 week, 2nd week, and 4th week using the PSQI sleep scale, while changes in resting-state functional connectivity (rsFC) were assessed at baseline as well as at the end of 4th week.

## Statistical analyses

A paired t-test was used to compare the differences in PSQI and EEG signals of whole brain 40HZ Gamma oscillation at baseline and after 2 or 4 weeks of 40Hz photoacoustic stimulation combined with blue light. For the rs-FC analysis, changes in mean functional connectivity at baseline and after the 4-week intervention were assessed using a paired t-test (one-sided), and a one-tailed p < 0.05 was considered as significant. The threshold for all other p values was chosen as 0.05. All statistical analyses were performed using R (Version 4.2).

### Functional Connectivity Analysis

#### MRI Scanning and Data Processing

All subjects were scanned using a 3.0 Tesla MRI scanner (GE Discovery 750). The three dimensions T1 weighted (3D T1) images were acquired using a Fast Spoiled Gradient Recalled sequence: echo time = 3.036 ms; repetition time = 7.336 ms; inversion time = 450 ms; flip angle = 11°; field of view = 260 × 260 mm^2^; matrix = 256 × 256; slice thickness = 1.2 mm; 196 sagittal slices. Rs-fMRI images were acquired using Gradient Recalled Echo—Echo Planar Imaging sequence: echo time = 30 ms; repetition time = 2,000 ms; flip angle = 77°; field of view = 240 × 240 mm^2^; matrix = 64 × 64; slice thickness = 4 mm; slice gap = 0 mm; 38 interleaved axial slices.

#### fMRI data preprocessing

Resting-state fMRI images were preprocessed using the Data Processing and Analysis for Resting-State Brain Imaging tools (DPABI, http://rfmri.org/dpabi) based on Statistical Parametric Mapping 12 (https://www.fil.ion.ucl.ac.uk/spm). The first 10 time points were not included in the analysis to consider scanner stabilization and the participants’adaptation to the environment. The remaining functional images were first corrected for within-can differ-rences in acquisition time between slices. The images were realigned to the middle volume to correct for interscan head motion. Subsequently, the processed images were registered to 3D T1 images and spatially normalized to a standard template (Montreal Neurological Institute). Thecorrected images were smoothed using a Gaussian kernel of 6×6×6 mm^3^ and then detrended. Nuisance covariates, including white matter, cerebrospinal fluid, and 24 motion parameters were regressed and temporal bandass filtering applied at a frequency range of 0.01– 0.1 Hz. Finally, subjects were excluded using the Jenkinson framewise displacement threshold of > 0.2 mm.

#### Functional connectivity between hippocampus and the nodes of default mode network (DM N)

We examined the intra-network functional connectivity of ROIs in the DMN, among medial prefrontal cortex (mPFC), posterior cingulate cortex (PCC), and precuneus (PCu), based on previous studies. Functional connectivity was measured using the DPABI toolbox. Parcellation of mPFC was based on the Human Connectome Project, and the parcellation of PCC and Pcu were performed based on AAL atlas. We also examined the functional connectivity between hippocampus and the ROIs of DMN (mPFC, PCC, and Pcu). Parcellation of hippocampus was based on the AAL atlas. The left and right mPFC, PCC, Pcu, and hippocampus were merged as signal ROIs.

#### Functional connectivity between hippocampus and whole brain regions

Processed fMRI data were subjected to further functional connectivity analysis. The AAL Atlas was used to extract the time courses and calculate the functional connectivity between right and hippocampus and the rest of brain regions (include 114 cortical, subcortical, and cerebellar regions).

#### Assessment of induced entrainment using EEG

EEG response was evaluated in 17 subjects to test whole brain 40Hz Gamma oscillation ratios at the same time under 5 stimulation modalities. The EEG data were pre-processed and analyzed using MNE-Python [1], using the Welch method (MNE-Python compute_psd function, method = ‘welch’, n_fft = 2048, window = Hamming) to extract the power spectral features of the EEG signals.

The oscillation ratio of an EEG channel (representing a specific brain region) was calculated based on the significant increase in the power spectral density of the oscillations generated by each channel at 40Hz relative to the surrounding 35Hz-45Hz frequency band. The strength of the EEG 40Hz Gamma oscillation response for each channel was categorized into significant response and very significant response, according to the parameters of the Shudan RWE data lake. The significant oscillation ratio of the whole brain was defined as the percentage of channels that reach a significant response while the highly significant ratio of the whole brain was defined as the percentage of channels that reach a highly significant response.

The thermogram of the brain was calculated based on median changes in the value of the 40Hz power spectral density (dB) generated by each channel in the stimulated state compared to the baseline resting state level.

## Result

### Baseline participant characteristics

The final four members included in the analysis were all female, as two male subjects withdrew from the study due to severe symptoms of COVID-19 and taking medication to improve insomnia, respectively. The median age of the population was 30 (IQR: 27.8-33.8), the median length of education was 12 years (IQR: 27.8-33.8) and the median PSQI score at baseline was 7.5 (IQR: 7-8.75) (**Table1**).

**Table1:**
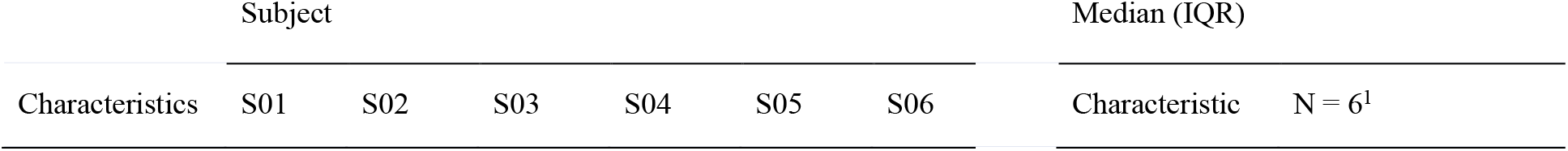
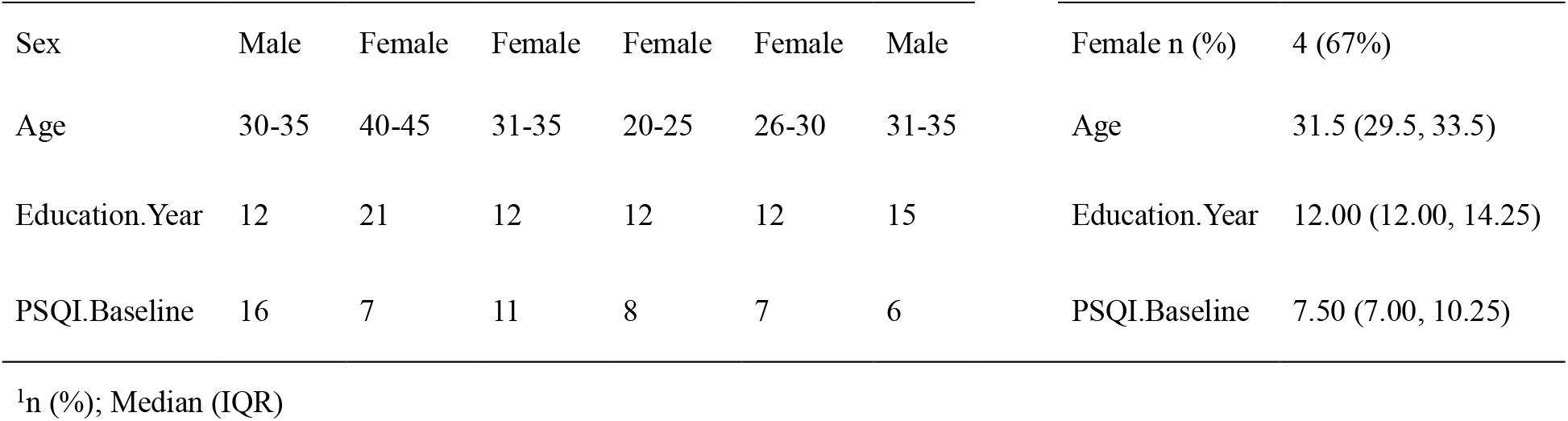
Baseline characteristics of the study participants

### EEG response for different stimulus modes

There were significant differences in the response ratios of brain regions among different 40 Hz photoacoustic stimulation modalities (p < 0.001, **Table 2**), with single stimulation modality (visual or audio single stimulation) producing lower 40 Hz Gamma oscillations than the combined stimulation modality consisting of visual and audio stimulation. The mean percentage of whole-brain 40Hz Gamma oscillations that reached significant levels after visual stimulation by looking at the front horizontally, visual stimulation by looking straight at the device, and audio stimulation were 38% (IQR: 25%-44%), 69% (IQR: 56%-75%) and 62% (IQR: 50%-88%), respectively, whereas the mean percentage of whole-brain 40Hz Gamma oscillations that reached significant levels after the combined stimulation modalities (visual stimulation by looking at the front horizontally+audio stimulation and visual stimulation by looking straight at the device+audio stimulation) were 75% (IQR: 62%-88%), and 75% (IQR: 62%-88%)(**Figure 2A**), respectively. The combined stimulation modality consisting of visual stimulation by looking straight at the device + audio stimulation generated the highest response rate suggesting that it was the strongest stimulation modality. However, the combined stimulation modality consisting of visual stimulation by looking at the front horizontally + audio stimulation was the optimal stimulation modality chosen for the subsequent 4-week stimulation. This is because under this stimulation modality, the subjects showed the same level of response rate as the strongest stimulation modality while looking at the front would help the subjects to better sustain long-term use of the device (as expected, we saw 0 exclusion due to being unable to stick with the daily use routine). In addition, the EEG signals showed that the combined stimulation modality not only generated greater whole brain response rate, but also had smaller variation than any single stimulation modality.

**Table2:** Whole-brain EEG response ratio among different stimulation modalities

| Characteristic | Response Ratio (Baseline) |
| --- | --- |
| Overall, N = 85<sup>1</sup> | 0.62 (0.44, 0.81) |
| Visual (looking at the front horizontally), N = 17<sup>1</sup> | 0.38 (0.25, 0.44) |
| Visual (looking straight), N = 17<sup>1</sup> | 0.69 (0.56, 0.75) |
| Auditory, N = 17<sup>1</sup> | 0.62 (0.50, 0.88) |
| Combined (looking at the front horizontally), N = 17<sup>1</sup> | 0.75 (0.62, 0.88) |
| Combined (looking straight), N = 17<sup>1</sup> | 0.75 (0.62, 0.88) |
| p-value<sup>2</sup> | <0.001 |

<sup>1</sup>Median (IQR); <sup>2</sup>Kruskal-Wallis rank sum test

**Figure 2.**
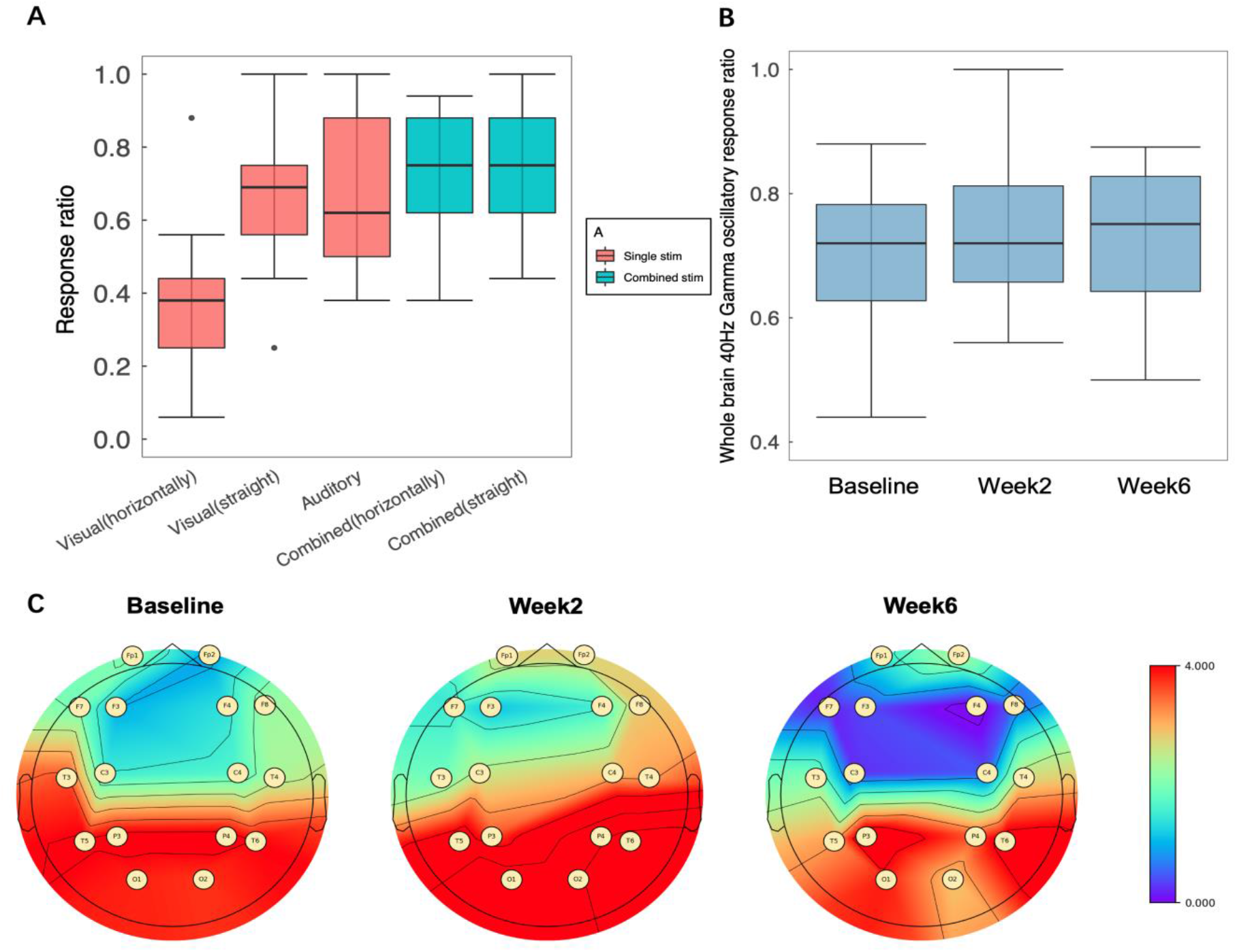
**(A)** Box plots of whole brain EEG response rate for different stimulation modalities, where blue represents combined stimulation modalities and red represents single stimulation modalities. The EEG response rate of significant 40HZ Gamma oscillation of combined stimulation was higher than that of single stimulation There was a significant difference between the two single visual stimulation modalities, but the difference between the two combined stimulation modalities was not significant. **(B)** Whole-brain EEG response rates at baseline, after 2-week of intervention, and 4-week of intervention (measured at the end of 6th week due to COVID-19) under the optimal stimulation modality. No signal decay was observed over time, demonstrating the stability of the long-term intervention. **(C)** Median values of intervention-induced changes in 40-Hz power spectral density (PSD, dB) for each channel at baseline, after 2-week intervention, and after 4-week intervention (measured at the end of 6th week due to COVID-19), showed an increase in the intensity of the 40-Hz Gamma oscillation response in the parietal, occipital, temporal, and frontal brain channels compared to the resting state.

### EEG response stability analysis

Four healthy subjects were expected to undergo EEG response stability testing after the 2nd and 4th week of the interventions, but due to the pandemic, the final EEG testing was actually tested at week 6, 2 weeks later than expected. There were no significant changes in the whole-brain response rate, and the intensity of the 40Hz Gamma oscillation response across different EEG channels.

The median percentage of channels with a significant response out of all channels was 72% (IQR: 62.75%-78.25%) at baseline, remained at 72% (IQR: 65.75%-81.25%) after 2 weeks of intervention, and had a median percentage of 75.1% (IQR: 64.25%-82.77%) after 6 weeks. The heat map (**Figure 2C**) showed an increase in the intensity of the 40Hz Gamma oscillatory response in the parietal, occipital, temporal, and frontal brain channels compared to the resting state. The 40Hz Gamma oscillatory response in the occipital lobe, which is the visual cortical centre, and in the temporal lobe, where the primary and secondary auditory cortices are located, were particularly strong.

### Sleep quality improvement

Sleep disturbances are more frequent and severe in patients with mild cognitive impairment (MCI) and AD compared to cognitively normal older adults. Accumulating clinical data suggests a strong bidirectional association between sleep disturbances and disease progression in AD, suggesting a vicious cycle contributing to AD progression (Bubu et al., 2020; Wang and Holtzman, 2020).

A total of 4 subjects completed the 4-week 40Hz photoacoustic stimulation combined with blue light intervention (**Table1**) and their sleep was assessed at baseline, the 2nd week and the 4th week using the PSQI sleep scale. The PSQI scale of behavioural data in healthy subjects demonstrated significant improvement in the sleep quality at the end of the 4-week intervention. All four subjects scored > 6 (median: 7.5) at baseline, an indication that they needed sleep quality improvement. After 4 weeks of the 40Hz photoacoustic stimulation combined with blue light intervention, there was a significant decrease in the PSQI scores (p = 0.028, **Figure 3**), with the median PSQI value decreasing from 7.5 at baseline to 5. A PSQI score ranging from 0 to 5 indicates that sleep quality was within an optimal level.

**Figure 3.**
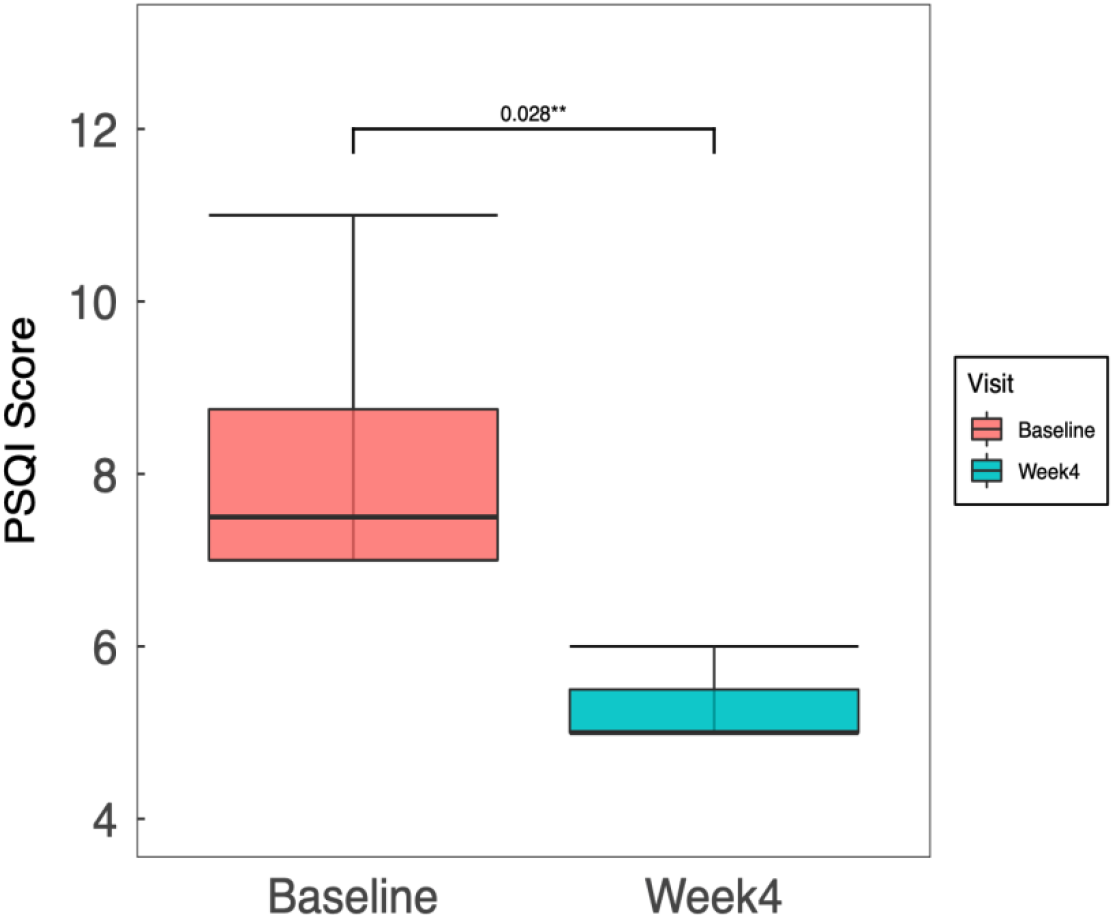
PSQI scores before and after the four-week intervention. The healthy subjects showed significant improvement in their sleep quality, with the median PSQI score decreasing from 7.5 at baseline to 5. Sleep quality was considered within optimal range when the PSQI score was less than 6.

### rs-FC result

After 4 weeks of intervention, resting-state fMRI of the brain showed significant increase in neural connectivity in 3 memory- and motor-related brain regions in the hippocampus and DMN regions **(Figure 4A)**. There was significant increase in functional connectivity after 4 weeks of intervention between the right hippocampus and the right medial superior frontal gyrus (one-sided paired t-test: 0.203, P = 0.01828), the left hippocampus and the right cerebellar area 7b (one-sided paired t-test: 0.203, P = 0.04527), the right hippocampus and the right angular gyrus (one-sided paired t-test: 0.203, P = 0.04527), and between the right hippocampus and the right angular gyrus (one-sided paired t-test: 0.229, P = 0.04059).

**Figure 4.**
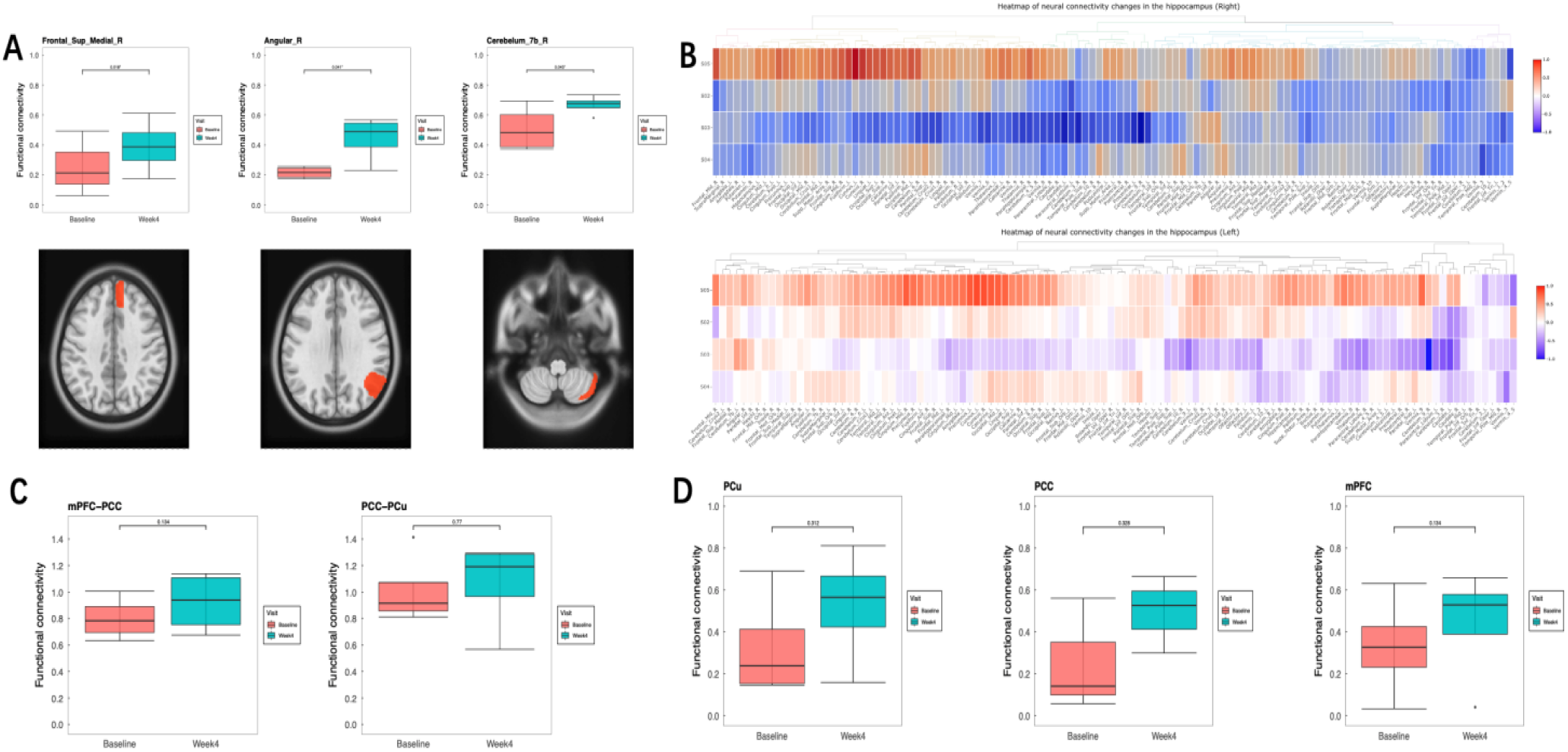
**(A)** Resting-state MRI data before and after four weeks of intervention showed significant enhancement of the hippocampus with the right superior medial frontal gyrus, the right cerebellar area 7b, and the right angular gyrus, p < 0.05. **(B)** Heat maps of neural connectivity changes between the hippocampus and the DMN region obtained from resting-state MRI data after four weeks of intervention showed a non-significant enhancement of the hippocampal region compared to that at baseline; **(C)** Functional connectivity changes between the posterior cingulate cortex (PCC) and precuneus (PCu), and between the medial prefrontal cortex (mPFC) and PCC, after four weeks of intervention. The median functional connectivity between PCC and PCu and between mPFC and PCC showed enhanced connectivity, as reflected by an increase in median number, before and after the four-week comparison, but the change was not significant. **(D)** The median functional connectivity between the hippocampus and PCC, PCu and mPFC all showed an increase after 4 weeks of intervention.

There was also increase in functional connectivity in the hippocampus and DMN regions, although not significant (**Figure 4B**), with a higher increase in the left hippocampus and DMN than in the right hippocampus after the 4-week intervention. The increase was probably due to the fact that the subjects were all right-handed and that the left side of the brain was more closely related to higher cognitive functions.

Functional connectivity between the hippocampal region and the posterior cingulate cortex (PCC) and precuneus (PCu) and between the PCC and medial prefrontal cortex (mPFC) was also elevated, although not significant (**Figure 4C**). Specifically, functional connectivity between the hippocampus and the mPFC was increased but not significant after the 4-week intervention (paired t-test: 0.110, 95% CI: (−0.060, 280), P = 0.1324). There was an increase in functional connectivity between the hippocampus and PCu (paired t-test; 0.197, 95% CI: (−0.320, 0.714), P = 0.312) and between the hippocampus and PCC (paired t-test; 0.162, 95% CI: -0.282, 0.608, P = 0.328) after the 4-week intervention, but the increase was not significant.

There was also an increase in functional connectivity between the PCC and mPFC (paired t-test: 0.12, 95% CI: (−0.068, 0.310), P = 0.1343), and between the PCC and PCu (paired t-test: 0.046, 95% CI: (−0.427, 0.521), P = 0.77) after the 4-week intervention, but the increase was not significant (**Figure 4D**). The increase was probably partly due to the fact that the healthy subjects had relatively higher baseline functional connectivity compared to those who were diagnosed with MCI or AD, and partly due to the limited intervention duration. The functional connection between PCC-PCu was better in the healthy population at baseline, but poor in patients with MCI/AD. In addition, the length of the intervention in this study was only 4 weeks, which may not have been sufficient to see a significant change. Current clinical trial studies have shown that the functional connection between PCC and PCu was not significant after 4 weeks, but was significantly enhanced when the length of intervention reached 8 weeks.

## Safety

Potential concerns about participant safety, compliance and tolerance to daily use of 40Hz photoacoustic stimulation combined with blue light was also assessed.

Safety was assessed using an adverse event questionnaire completed on site by the subjects each week. Adherence was assessed by asking the subjects to receive the intervention at the same time and in the same location, with a staff member recording the start and end times of the intervention on site and monitoring the implementation of the intervention on site.

Serious adverse events associated with 40Hz photoacoustic stimulation, such as including dizziness, tinnitus, headache and worsening hearing loss, were not reported. There were reports of mild adverse events such as dry eyes (N=2) and drowsiness (N=1). The symptoms of individuals who reported dry eyes improved after 1 week of intervention and the symptoms did not recur with continued intervention. The subject who reported drowsiness was excluded from the study, as he fell asleep during the intervention and was unable to complete a full EEG response test. Evaluation of adverse events was to be carried out for upto 12 weeks, which may still be ongoing at the time of publication of this report.

## Discussion

The aim of this study was to investigate the effect of 40Hz photoacoustic stimulation combined with blue light in improving brain function and sleep quality in a healthy population. We first compared EEG response under 5 different stimulation modalities to identify the optimum stimulation modality. We then assessed the stability and consistency of the optimum stimulation modality by recording EEG changes in whole brain gamma oscillatory response at different time points over 1 month of the intervention. We also determined the effect of the intervention on sleep improvement in healthy individuals with sleep issues by recording Pittsburgh Sleepiness Scale (PSQI) scores at different time points during the intervention. We observed an increase in strength of functional connections in various relevant areas of the brain.

Combined stimulation produced greater whole-brain response than single-source stimulation based on results of EEG response rate. In addition, the whole-brain response rate to 40Hz Gamma oscillation and the intensity of the EEG response in each channel did not change significantly for all subjects over the 4-week intervention, suggesting that the combined stimulation produced effective and stable Gamma oscillations across all the subjects.

Blue light effectively regulates circadian rhythms (Boutrel B, 2004) and improves subjective sleep quality and daytime functioning the next day in healthy subjects (Li D, 2022). In our study, we observed a significant decrease in the overall PSQI score after 4 weeks of 40Hz photoacoustic stimulation combined with blue light, indicating that the combined stimulation significantly improved the sleep quality of the subjects within one month.

The default network (DMN) consists mainly of areas such as the posterior cingulate gyrus (PCC), medial prefrontal cortex (mPFC), precuneus and inferior parietal lobule. There are connections to the medial temporal lobe system such as the hippocampus and parahippocampal gyrus. If functional connectivity between the hippocampus and the default mode network (DMN) is enhanced, this change is often thought to facilitate enhanced cognitive functions such as sensory processing, attention, and memory processing (Mak LE, 2017). A study conducted in 2018 reported improved functional connectivity between the hippocampus and DMN in AD patients as symptoms improved (Chao, L. L. 2019). The improvement in symptoms was associated with an increase in network connectivity.

Previously, studies focused on the use of photoacoustic stimulation to alleviate cognitive decline in AD and MCI patients. Therefore, few investigations have been conducted to explore the preventive effects of photoacoustic stimulation in healthy populations. In this study, we investigated the effect of 40HZ photo acoustic stimulation combined with blue light on brain activity patterns in a healthy population. We found that after 4 weeks of the intervention, the resting-state functional MRI of the brain showed enhanced functional connectivity in the hippocampal and DMN regions. Particularly, there were significant differences in functional connectivity between the right hippocampus and the right medial superior frontal gyrus, the left hippocampus and the right cerebellar area 7b, and the right hippocampus and the right angular gyrus. In addition, we found varying degrees of enhancement between the hippocampus and the posterior cingulate cortex (PCC) and precuneus (Pcu) and between the PCC and medial prefrontal cortex (mPFC), although the differences were not significant, which was consistent with the results of previous clinical trials (Chan D, 2022). We speculate that this observation may be related to the short duration of the intervention and the fact that all participants were healthy and had stronger baseline connectivity. Because of the ceiling effect in healthy young adults; that is, the relatively small improvement in cognitive performance (Tort, A. B. L., 2009), we did not perform cognitive behavioral performance assessment. Therefore, significant changes in wider brain regions and improvements in cognitive performance, if any, may require longer intervention periods, repeated intervention cycles or higher intervention intensities (Phipps-Nelson, 2003).

All participants showed good tolerance to the stimulation and adhered to the intervention satisfactorily. However, 2 participants were removed from the group due to severe COVID-19 symptoms and taking medication to improve insomnia, respectively. Some participants experienced discomfort with dry eyes and astringent eyes during week 1 of the intervention, but the discomfort disappeared in week 2 and no serious adverse events associated with the intervention were observed throughout the study.

There are several limitations to this study. To begin with, the small sample size and short duration of the intervention may have limited the statistical power and magnified individual differences in responses to the intervention, leading to overestimation and low reproducibility of the results. Therefore, further studies with larger sample size are needed to confirm the consistent improvement of sleep quality and brain function in healthy populations. Although there was heterogeneity among the participants, we made corrections where possible. For instance, participants received the same intensity and form of light-sound intervention at the same place at the same time each day. To ensure the reliability of the various connectivity results shown at the group level, all functional MRI experiments were completed at similar time points using the same MRI testing equipment.

Secondly, we only established our pre- and post-control groups, but no control groups such as a placebo test group, or a test subgroup set up for either the blue light or 40Hz photoacoustic intervention, making it difficult to clearly separate the individual effects caused by 40HZ photo acoustic intervention from those caused by blue light. Although the combined light intervention could be considered as a”combo set” for convenient use in daily life which allows users to switch between the 40HZ white light and blue light in one device, there is a need to explore the mechanisms of the two lights as well as whether there is a placebo effect triggered by sham light intervention. Currently, we know that sham stimulation may have unintended effects in a single stimulus (Lin Z, 2021). Whereas additional activation in bilateral hippocampal regions in the object recognition task was observed only in the 40Hz light stimulation group, additional prefrontal cortex (prefrontal cortex PFC), which is known for object discrimination, showed additional activation in the sham stimulation group.

Despite these limitations, we present preliminary evidence that the 40Hz photoacoustic stimulation combined with blue light significantly reduced PSQI scores for all participants, indicating significant improvement in sleep quality. In addition, we observed enhanced functional connectivity in the hippocampus under the default mode network in resting-state functional magnetic resonance. These findings demonstrate that 40Hz photoacoustic stimulation combined with blue light may be a new, non-invasive, non-pharmacological approach to improve brain functions and sleep quality in healthy populations. Further testing in larger sample sizes, longer treatment durations and a blinded, placebo-controlled design is needed to examine the consistency of this novel treatment approach in improving brain function, sleep quality and other behavioral performance in healthy populations.

## Data Availability

All data produced in the present study are available upon reasonable request to the authors.

## Supplements

**Appendix S1:**
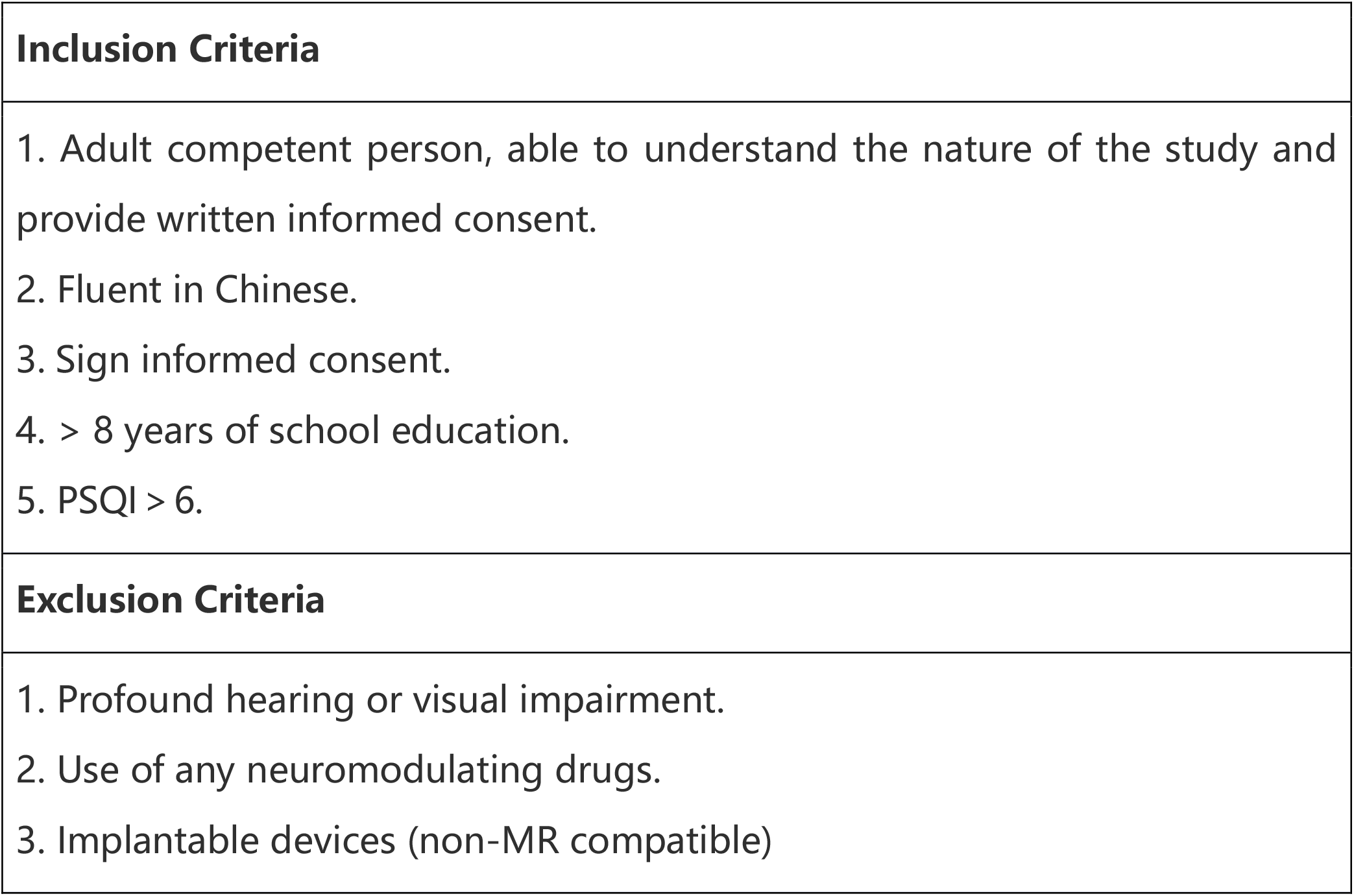
Inclusion/Exclusion Criteria

| Inclusion Criteria |
| --- |
| 1. Adult competent person, able to understand the nature of the study and provide written informed consent.<br>2. Fluent in Chinese.<br>3. Sign informed consent.<br>4. > 8 years of school education.<br>5. PSQI > 6. |
| Exclusion Criteria |
| 1. Profound hearing or visual impairment.<br>2. Use of any neuromodulating drugs.<br>3. Implantable devices (non-MR compatible) |

## References

1. Wang Y, Pan Y, Li H. What is brain health and why is it important? BMJ. 2020 Oct 9;371:m3683. doi: 10.1136/bmj.m3683.

2. GBD 2016 Neurology Collaborators. Global, regional, and national burden of neurological disorders, 1990-2016: a systematic analysis for the Global Burden of Disease Study 2016. Lancet Neurol 2019;18:459-80. doi:10.1016/S1474-4422(18)30499-X

3. Jia J, Wei C, Chen S, et al. The cost of Alzheimer’s disease in China and re-estimation of costs worldwide. Alzheimers Dement. 2018 Apr;14(4):483–491. doi: 10.1016/j.jalz.2017.12.006.

4. World Alzheimer Report. The state of the art of dementia research: New frontiers[R]. London. Alzheimer’s Disease International. 2018,

5. World Alzheimer Report 2018 | Alzheimer’s Disease International (ADI) (alzint.org)

6. Jia L, Du Y, Chu L, et al. Prevalence, risk factors, and management of dementia and mild cognitive impairment in adults aged 60 years or older in China: a crosssectional study[J]. Lancet Public Health, 2020, 5(12): e661e671.DOI: 10.1016/s24682667(20)301857.

7. Tisher A, Salardini A. A Comprehensive Update on Treatment of Dementia. Semin Neurol. 2019 Apr;39(2):167–178. doi: 10.1055/s-0039-1683408. Epub 2019 Mar 29. PMID: 30925610.

8. World Health Organization. Global action plan on the public health response to dementia 2017-2025. Available: https://www.who.int/publications/i/item/9789241513487 [Accessed 14 Dec 2021].

9. Girardeau G, Lopes-Dos-Santos V. Brain neural patterns and the memory function of sleep. Science. 2021 Oct 29;374(6567):560–564. doi: 10.1126/science.abi8370.

10. Soldatos CR, Allaert FA, Ohta T, Dikeos DG. How do individuals sleep around the world? Results from a single-day survey in ten countries. Sleep Med. 2005 Jan;6(1):5–13. doi: 10.1016/j.sleep.2004.10.006.

11. Xu W, Tan L, Wang HF, et al. Meta-analysis of modifiable risk factors for Alzheimer’s disease. J Neurol Neurosurg Psychiatry. 2015 Dec;86(12):1299–306. doi: 10.1136/jnnp-2015-310548.

12. Pan Y, Li H, Wardlaw JM, Wang Y. A new dawn of preventing dementia by preventing cerebrovascular diseases. BMJ. 2020 Oct 9;371:m3692. doi: 10.1136/bmj.m3692.

13. Chang CH, Lane HY, Lin CH. Brain Stimulation in Alzheimer’s Disease. Front Psychiatry. 2018 May 22;9:201. doi: 10.3389/fpsyt.2018.00201.

14. Sabbagh M, Sadowsky C, Tousi B, et al. Effects of a combined transcranial magnetic stimulation (TMS) and cognitive training intervention in patients with Alzheimer’s disease. Alzheimers Dement. 2020 Apr;16(4):641–650. doi: 10.1016/j.jalz.2019.08.197.

15. Gaitán JM, Moon HY, Stremlau M, et al. Effects of Aerobic Exercise Training on Systemic Biomarkers and Cognition in Late Middle-Aged Adults at Risk for Alzheimer’s Disease. Front Endocrinol (Lausanne). 2021 May 20;12:660181. doi: 10.3389/fendo.2021.660181.

16. Laccarino HF, Singer AC, Martorell AJ, et al. Gamma frequency entrainment attenuates amyloid load and modifies microglia. Nature. 2016 Dec 7;540(7632):230–235. doi: 10.1038/nature20587.

17. Adaikkan C, Middleton SJ, Marco A, et al. Gamma Entrainment Binds Higher-Order Brain Regions and Offers Neuroprotection. Neuron. 2019 Jun 5;102(5):929-943.e8. doi: 10.1016/j.neuron.2019.04.011.

18. Boutrel B, Koob GF. What keeps us awake: the neuropharmacology of stimulants and wakefulness promoting medications. Sleep. 2004;27(6):1181–1194. doi:10.1093/sleep/27.6.1181

19. Li D, Fang P, Liu H, et al. The Clinical Effect of Blue Light Therapy on Patients with Delayed Sleep-Wake Phase Disorder. Nat Sci Sleep. 2022 Jan 18;14:75–82. doi: 10.2147/NSS.S344616.

20. Mak LE, Minuzzi L, MacQueen G, et al. The Default Mode Network in Healthy Individuals: A Systematic Review and Meta-Analysis. Brain Connect. 2017 Feb;7(1):25–33. doi: 10.1089/brain.2016.0438.

21. Greicius MD, Srivastava G, Reiss AL, et al. Default-mode network activity distinguishes Alzheimer’s disease from healthy aging: evidence from functional MRI[J]. Proc Natl Acad Sci USA, 2004, 101(13): 4637–4642. DOI:10.1073/pnas.0308627101.

22. Zhong YF, Huang LY, Cai SP, et al. Altered effective connectivity patterns of the default mode network in Alzheimer’s disease: an fMRI study[J]. Neurosci Lett, 2014, 578: 171–175. DOI:10.1016/j.neulet.2014.06.043.

23. Chao, L.L. (2019). Effects of home photobiomodulation treatments on cognitive and behavioral function, cerebral perfusion, and resting-state functional connectivity in patients with dementia: a pilot trial. Photobiomodul. Photomed. Laser Surg. 37, 133–141. doi: 10.1089/photob.2018.4555

24. He Q, Colon-Motas KM, Pybus AF, et al. A feasibility trial of gamma sensory flicker for patients with prodromal Alzheimer’s disease. Alzheimers Dement (N Y). 2021 May 13;7(1):e12178. doi: 10.1002/trc2.12178.

25. Cimenser A, Hempel E, Travers T, et al. Sensory-Evoked 40-Hz Gamma Oscillation Improves Sleep and Daily Living Activities in Alzheimer’s Disease Patients. Front Syst Neurosci. 2021 Sep 24;15:746859. doi: 10.3389/fnsys.2021.746859.

26. Barth AM, Mody I. Changes in hippocampal neuronal activity during and after unilateral selective hippocampal ischemia in vivo. J Neurosci. 2011 Jan 19;31(3):851–60. doi: 10.1523/JNEUROSCI.5080-10.2011.

27. Jones, M., McDermott, B., Oliveira, B. L., et al. (2019). Gamma band light stimulation in human case studies: groundwork or potential Alzheimer’s disease treatment. J. Alzheimers Dis. 70, 171–185. doi: 10.3233/JAD-190299.

28. Yao, Y., Zhang, W., Ming, R., et al. (2020). Noninvasive 40-Hz light flflicker rescues circadian behavior and abnormal lipid metabolism induced by acute ethanol exposure via improving SIRT1 and the circadian clock in the liver-brain axis. Front. Pharmacol. 11:355. doi: 10.3389/fphar.2020.00355.

29. Tseng, P., Chang, Y. T., Liang, W. K., et al. (2016). The critical role of phase difffference in gamma oscillation within the temporoparietal network for binding visual working memory. Sci. Rep. 6:32138. doi: 10.1038/srep32138.

30. Tort, A. B. L., Komorowski, R. W., Manns, J. R., Kopell, N. J., and Eichen-Baum, H. (2009). Theta–gamma coupling increases during the learning of item–context associations. Proc. Natl. Acad. Sci. U.S.A. 106, 20942–20947. doi: 10.1073/pnas.0911331106

31. Phipps-Nelson, J., Redman, J. R., Dijk, D. J., and Rajaratnam, S. M. (2003). Daytime exposure to bright light, as compared to dim light, decreases sleepiness and improves psychomotor vigilance performance. Sleep 26, 695–700. doi: 10.1093/sleep/26.6.695

32. Lin Z, Hou G, Yao Y, Zhou Z, Zhu F, Liu L, Zeng L, Yang Y, Ma J. 40-Hz Blue Light Changes Hippocampal Activation and Functional Connectivity Underlying Recognition Memory. Front Hum Neurosci. 2021 Dec 16;15:739333. doi: 10.3389/fnhum.2021.739333.

33. Chan, Diane et al. “Gamma frequency sensory stimulation in mild probable Alzheimer’s dementia patients: Results of feasibility and pilot studies.” PloS one vol. 17, 12 e0278412. 1 Dec. 2022, doi:10.1371/journal.pone.0278412

